# Early B-cell transcription factor-2 defect as a novel cause of lipodystrophy: disruption of the adipose tissue character and integrity

**DOI:** 10.1101/2024.06.24.24309093

**Authors:** Maria C. Foss-Freitas, Donatella Gilio, Andre Monteiro da Rocha, Lynn Pais, Melanie C. O’Leary, Heidi L. Rehm, Adam Neidert, Miriam S. Udler, Patrick Seale, Elif A. Oral, Tae-Hwa Chun

## Abstract

We report a novel cause of partial lipodystrophy associated with early B cell factor 2 (*EBF2*) nonsense variant (*EBF2* 8:26033143 C>A, c.493G>T, p.E165X) in a patient with an atypical form of partial lipodystrophy. The patient presented with progressive adipose tissue loss and metabolic deterioration at pre-pubertal age. *In vitro* and *in vivo* disease modeling demonstrates that the *EBF2* variant impairs adipogenesis, causing excess accumulation of undifferentiated CD34^+^ cells, extracellular matrix proteins, and inflammatory myeloid cells in subcutaneous adipose tissues. Thus, this *EBF2* p.E165X variant disrupts adipose tissue structure and function, leading to the development of partial lipodystrophy syndrome.

## Main Text

The hallmark of lipodystrophy syndromes (LD) is loss of adipose tissue (AT) and resulting deficiency in function, leading to insulin resistance and metabolic syndrome^1^. These diseases can present as generalized or partial LD^2^. The genetic underpinnings of about 30% of generalized LD and about 50% of partial lipodystrophy (PLD) syndromes remain unsolved^3^. To better define the genetic causes of LD, we initiated genetic screenings of individuals affected by LD using whole-genome sequencing (WGS) and next-generation sequencing panels. With these efforts, we are encountering an increasing number of gene variants of unknown significance (VUS)^4^; however, defining the biological roles played by these VUS in the pathogenesis of LD remains crucial. Through these efforts, we identified a novel heterozygous nonsense variant of the early B-cell factor 2 (*EBF2*), encoding a helix-loop-helix transcription factor, in a patient with atypical PLD. Murine *Ebf2* plays a crucial role in mesenchymal tissue development, including adipocyte differentiation, brown fat development, and bone metabolism^5-10^. However, the role of *EBF2* in human AT development is unknown. A subset of white adipocytes of human visceral AT expresses *EBF2* ^11^, and an *EBF2* single nucleotide polymorphism is associated with waist-hip ratio^11,12^, implicating the potential role of *EBF2* in regulating human AT function.

Therefore, there was biological plausibility in investigating the impact of the newly identified nonsense *EBF2* variant to determine if it played a causal role in our patient’s presentation. Here, we describe the functional impact of the observed *EBF2* nonsense variant on AT development and function through *in vitro* and *in vivo* disease modeling experiments.

## Methods

(Details and statistical methods are in the supplementary appendix.)

### Patient studies

Our team has followed this patient since pre-pubertal age, and she has participated in research studies that allowed clinical phenotyping and tissue biopsies^13,14^. After informed consent was obtained, an experienced plastic surgeon obtained AT biopsies from the back of the neck, abdomen, and upper thigh regions of the patient.

### DNA sequencing

WGS and data processing were performed using the Genomics Platform at the Broad Institute of MIT and Harvard.

### *In vitro* disease modeling

3T3-L1 mouse preadipocytes and primary human preadipocytes were used for *in vitro* disease modeling. Multiple siRNA oligos and lentiviral shRNA constructs targeting *Ebf2* were used for transient and permanent gene silencing. Wild-type and truncated variants of *EBF2* were expressed using bicistronic lentiviral constructs to assess the functional difference between the wild-type and the nonsense variant of *EBF2*. An *EBF2* reporter assay was performed to determine EBF2 transcriptional activity. Total RNA was extracted from cells and analyzed using targeted real-time qPCR and bulk RNA sequencing.

### *In vivo* disease modeling

The University of Michigan Transgenic Mouse Core generated a knock-in mouse model by inserting the exact single nucleotide variant, *Ebf2* p.E165X, using Crispr-Cas technology. The mice were backcrossed into the C57BL/6J strain for five generations before analysis.

## Results

### Case Report

The patient’s complex clinical presentation has been reported previously^14 13^. She is a young woman with PLD who first presented with progressive AT loss affecting hips and legs (Fig. 1A,B). No lipodystrophy phenotype was observed in her relatives. One of her parent, diagnosed with atypical lupus, heart disease, and type 2 diabetes, died prematurely at a young age; therefore, we do not have access to the genetic material. The patient also displayed scoliosis, hand contracture, and hypogonadotropic hypogonadism^13^. During follow-up, she developed progressive liver enzyme elevations and proteinuria and underwent liver and kidney biopsies. Her liver showed hepatocyte ballooning with excess lipid accumulation and fibrosis; her kidney showed fibrotic extracellular matrix (ECM) accumulation in the glomerulus and interstitial space (Supplement-Fig. 1), which was called “Alport-like pathology” by the case pathologist. A Clinical Laboratory Improvement Amendments (CLIA) certified lipodystrophy gene panel (*AGPAT2, AKT2, BSCL2, CAV1, CIDEC, LMNA, PLIN1, PPARG, PTRF, TBC1D4* and *ZMPSTE24*)^13^ did not detect any known variants. Subsequently, we performed WGS on samples from the proband, mother, and two siblings. We identified a heterozygous nonsense gene variant of zinc-knuckle DNA-binding domain containing transcription factor, *EBF2*, which we confirmed with targeted Sanger sequencing (Fig. 1C). The heterozygous nonsense variant of *EBF2* (NM_022659.4) in exon 6, leads to the premature termination of EBF2 at amino acid position 165 (EBF2 8:26033143 C>A, c.493G>T, p.E165X).

**Figure 1:**
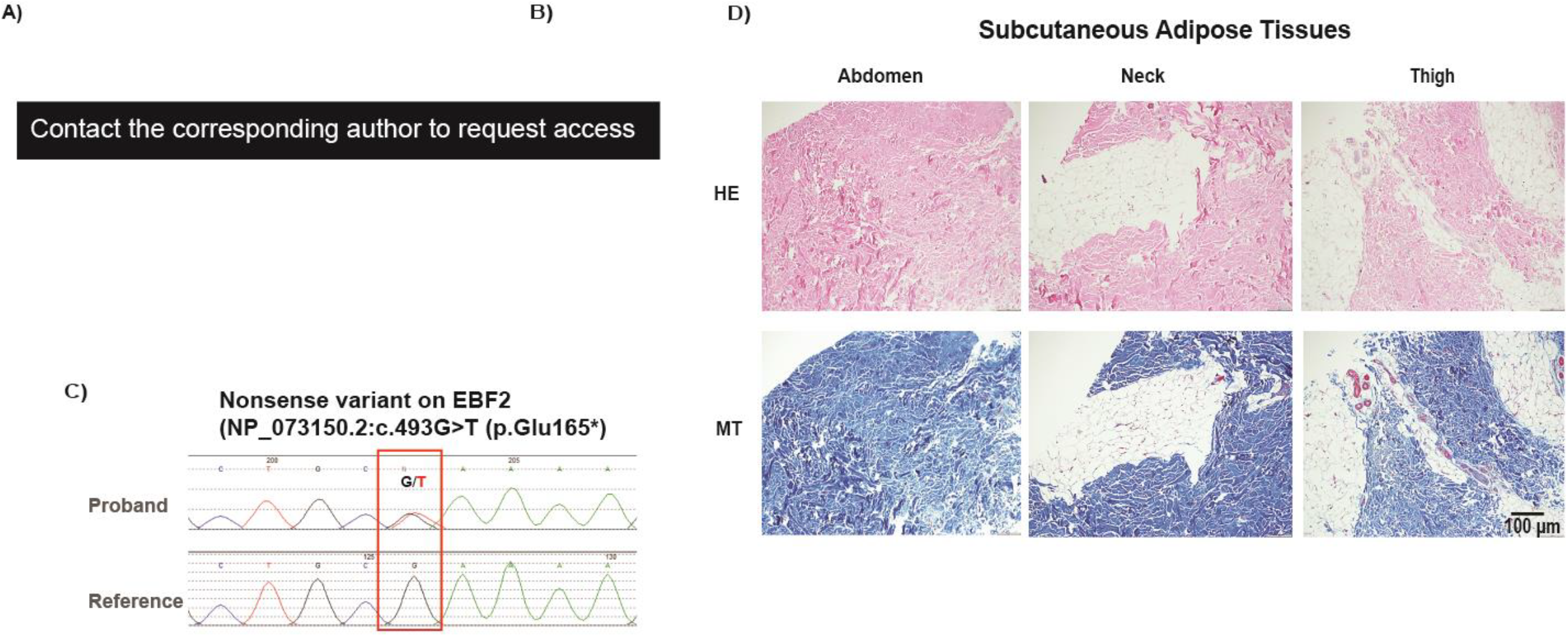
Progressive partial loss of AT in a patient with atypical partial lipodystrophy. A) Patient picture showing body fat distribution in front (far left) and side (second from left) panels, increased dorsal neck fat pad and acanthosis nigricans (arrow) (second from the right, upper), prominent superficial veins (arrows) due to fat loss in the legs (second from the right, lower). B) “Fat shadow” was obtained from the DEXA scan showing truncal AT distribution. C) Sanger sequence demonstrates the G>T nucleotide change in proband and reference sequence in non-affected mother. D) Hematoxylin-eosin (HE) and Masson Trichrome (MT) staining of subcutaneous AT (SQ) of the patient from different AT depots (abdomen, neck and thigh), showing AT surrounded by excess collagen deposition.

Biopsies of subcutaneous adipose tissues at three different fat depots (neck, abdomen, hip) showed increased ECM accumulation and inflammation (Fig. 1D), suggesting that disruption of tissue integrity may underlie this patient’s metabolic dysfunction.

### *EBF2* p.E165X *variant impairs adipocyte differentiation and function*

We knocked down the endogenous *Ebf2* gene in 3T3-L1 preadipocytes using two siRNA oligonucleotides (Supplemental-Fig. 2A-B). *Ebf2* silencing significantly impaired adipogenesis, as shown with reduced lipid accumulation and gene expression of adipocyte genes, *Pparg* and *Fabp4* (Supplemental-Fig. 2C). To perform a rescue experiment to assess the adipogenic potential of *EBF2*, we used five different lentiviral shRNA constructs to permanently knockdown *Ebf2* expression in 3T3-L1 cells and selected a clone (clone-4) that showed the specific suppression of *Ebf2* relative to *Ebf1* (Supplemental-Fig. 2D). When we reconstituted *Ebf2*-silenced 3T3-L1 cells with *EBF2* constructs using lentiviral gene transfer, full-length *EBF2* (*EBF2-full*) restored robust adipogenesis with enhanced expression of *Pparg* and *Fabp4* (Fig. 2A-C); however, the restoration of adipogenesis was not observed with the nonsense *EBF2* variant (p.E165X, *EBF2-mut*). Adipogenic potential conferred by the lentiviral *EBF2* gene paralleled the transcriptional activity of EBF2 as determined by reporter assays (Fig. 2D). When unmodified 3T3-L1 cells (which maintain endogenous *Ebf2* expression) (Fig. 2E) and human preadipocytes (which express *EBF2*) (Supplemental-Fig. 3A) were transduced with the *EBF2-mut*, these cells displayed significantly impaired adipogenesis, as shown by decreased lipid droplet content and reduced expression of *PPARG* and *FABP4*. These results suggest a loss of function and a potentially dominant-negative effect exerted by the truncated *EBF2* nonsense variant (Fig. 2F-H and Supplemental-Fig. 3B-E). We then performed bulk RNA sequencing of human preadipocytes, transduced with the lentivirus constructs of control, *EBF2-full*, and *EBF2-mut*, and stimulated with adipogenic cocktails (Fig. 2I). The gene transfer of *EBF2-full* and *EBF2-mut* led to differential gene expression in the ECM and cytokine-cytokine receptor interactions pathways (Fig. 2J-K). The *EBF2*-*mut* repressed the expression of *COL1A1, COL1A2, COL4A1, THBS1*, and *TNXB* while increasing the expression of *LAMB3, SPP1, LAMA1, ITGA10, ITGA5* (Fig. 2J). The *EBF2-mut* also increased the expression several cytokines and growth factors, including *IL1A, IL1B, CXCL2, CCL5, IL24*, and *TGFB2* (Fig. 2K). Together, these results reveal that the nonsense *EBF2* variant not only acts as a loss-of-function variant but partly exerts a dominant negative effect on adipogenesis and gene expression in the pathways of ECM remodeling and cytokine-cytokine receptor interaction. Consistent with these *in vitro* findings, the biopsied AT from our patient showed dysmorphic adipocytes surrounded by abundant ECM proteins and eosinophilic amorphous structures with excess myeloid cell infiltration (Fig. 1D).

**Fig. 2.**
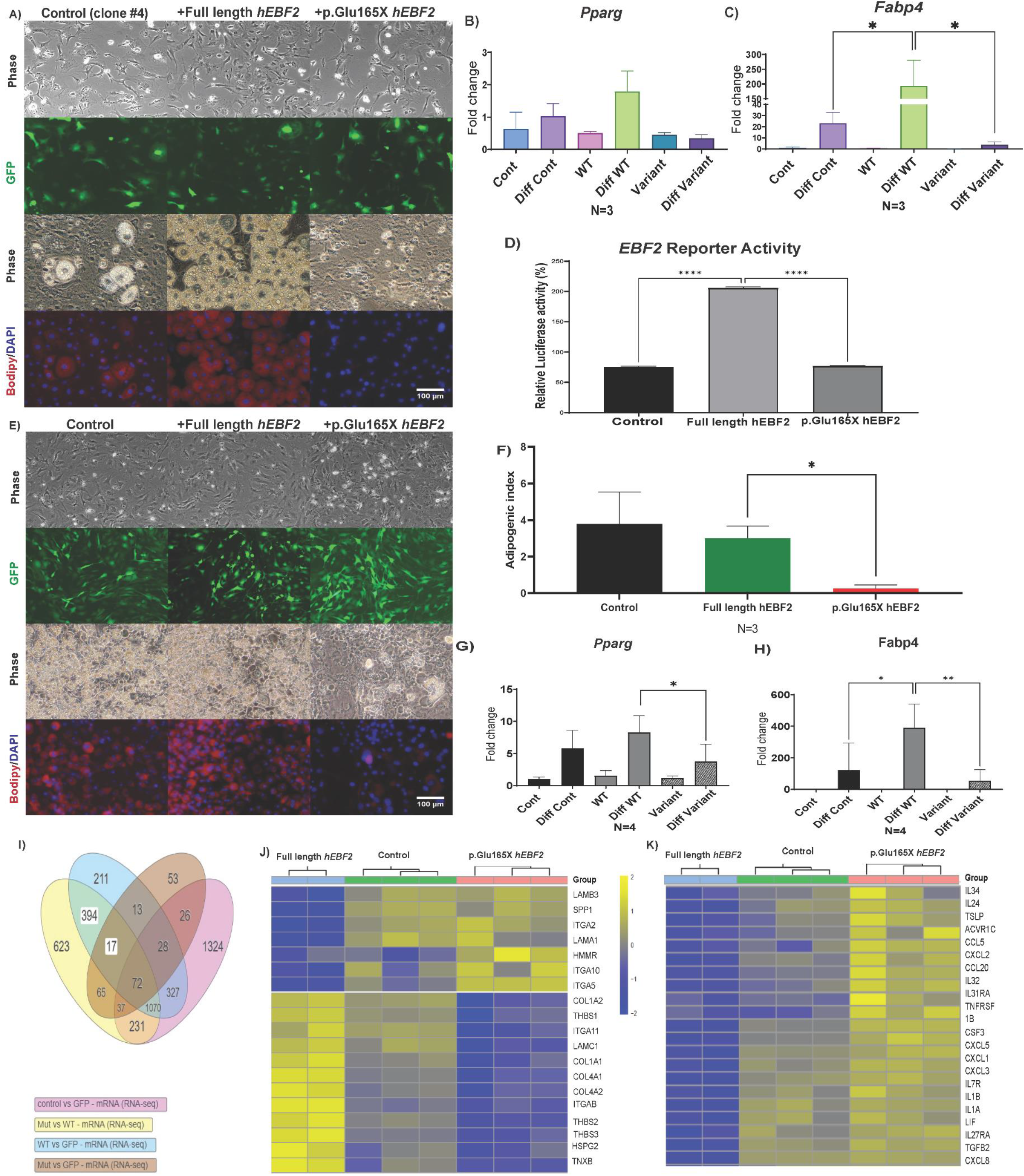
In vitro studies to determine the effect of *EBF2* nonsense variant in adipogenesis. A) Ebf2-silenced 3T3-L1 cells transduced with bicistronic empty GFP, EBF2-full GFP, and EBF2-mut GFP and adipogenesis induced. Lipid stained with BODIPY (red), nuclei (DAPI, blue), and GFP (green) (n=3). A representative figure is shown. Repeated more than four times. B) Fabp4 expression, and C) Pparg expression RT-qPCR (Diff= adipogenesis induced, data represents Mean±SEM, n=3, *p=0.02, statistical test performed by One way ANOVA with Bonferroni’s multiple comparisons). D) E-box reporter assay. HEK cells transfected EBF luc reporter (data represents Mean±SEM, n=5, ***p<0.001, representative data, statistical test performed by One way ANOVA with Tukey’s multiple comparisons). E) Mouse 3T3-L1 preadipocytes transduced with lentivirus control, EBF2-full, and EBF2-mut. Adipogenesis was induced and lipid droplet accumulation was assessed (BODIPY= red, nucleus= blue) and GFP (green). A representative figure is shown. Repeated more than four times. Adipocytes were quantified using Image J software with ADIPOQ plug-in. Cells transduced with lentiviral EBF2-mut show less adipogenic index (data represents Mean±SEM, n=3, *p=0.01, statistical test performed by One way ANOVA with Tukey’s multiple comparisons). G) *Pparg* and (H) *FABP4* gene expression from mouse 3T3-L1 preadipocytes transduced with lentivirus control, EBF2-full, and EBF2-mut (Diff= adipogenesis induced, data represents Mean±SEM, n=4, *p=0.03, **p=0.01, statistical test performed by One way ANOVA with Tukey’s multiple comparisons). I) Venn diagram of bulk RNA-seq of human SQ preadipocytes differentiated after lentiviral transduction of *EBF2-full, control*, and *EBF2-mut*. A cohort of ECM genes (left) (J) and cytokine-cytokine receptor genes (right) (K) are differentially regulated by EBF2 compared to the control.

### Knock-in mouse model phenocopies pleiotropic effects of the nonsense EBF2 variant on tissue structure and function

Based on these data, we hypothesize that the nonsense *EBF2* variant underlies the fibrotic and inflammatory AT damage observed in this patient. To test the hypothesis, we generated a knock-in mouse model (Fig. 3A). While we had no difficulty obtaining heterozygous (HET) mice from HET to C57BL/6J mice breeding, we could rarely obtain homozygous (HM) knock-in mice, raising concerns about perinatal lethality. A surviving female HM mouse lacked perigonadal WAT and showed rudimentary subcutaneous and inguinal WAT with dysmorphic adipocytes and extensive accumulation of eosinophilic fibrillar structures (Fig. 3B), which phenocopies the subcutaneous adipose tissues of the patient (HUM for human) displaying the limited number of adipocytes surrounded by excess ECM proteins (Fig. 3C). HET mice similarly demonstrated fibrotic tissue alteration in subcutaneous AT (Fig. 3B). Imaging cytometry (CyTOF) confirmed the deposition of type 1 collagen and the increased number of CD34^+^ cells and macrophages (F4/80^+^ cells in mice and CD68^+^ cells in the patient) (Fig. 3D). CyTOF unveiled the infiltration of neutrophils (Ly6G^+^ cells in mice and CD15^+^ cells in humans) and monocytes (CD11b^+^ cells in mice and CD11c^+^ cells in humans) in HM and HUM tissues, respectively (Supplemental-Fig. 4A). Immunostaining further validated these findings, showing the reduced number of PLIN1^+^ adipocytes and increased deposition of type 1 collagen, fibronectin, and elastin (Fig. 3E) coupled with an increased number of smooth muscle cell actin (SMA)^+^ cells, underscoring fibrotic phenotype of AT in the patient and KI mouse model (Supplemental-Fig. 4B).

**Fig. 3.**
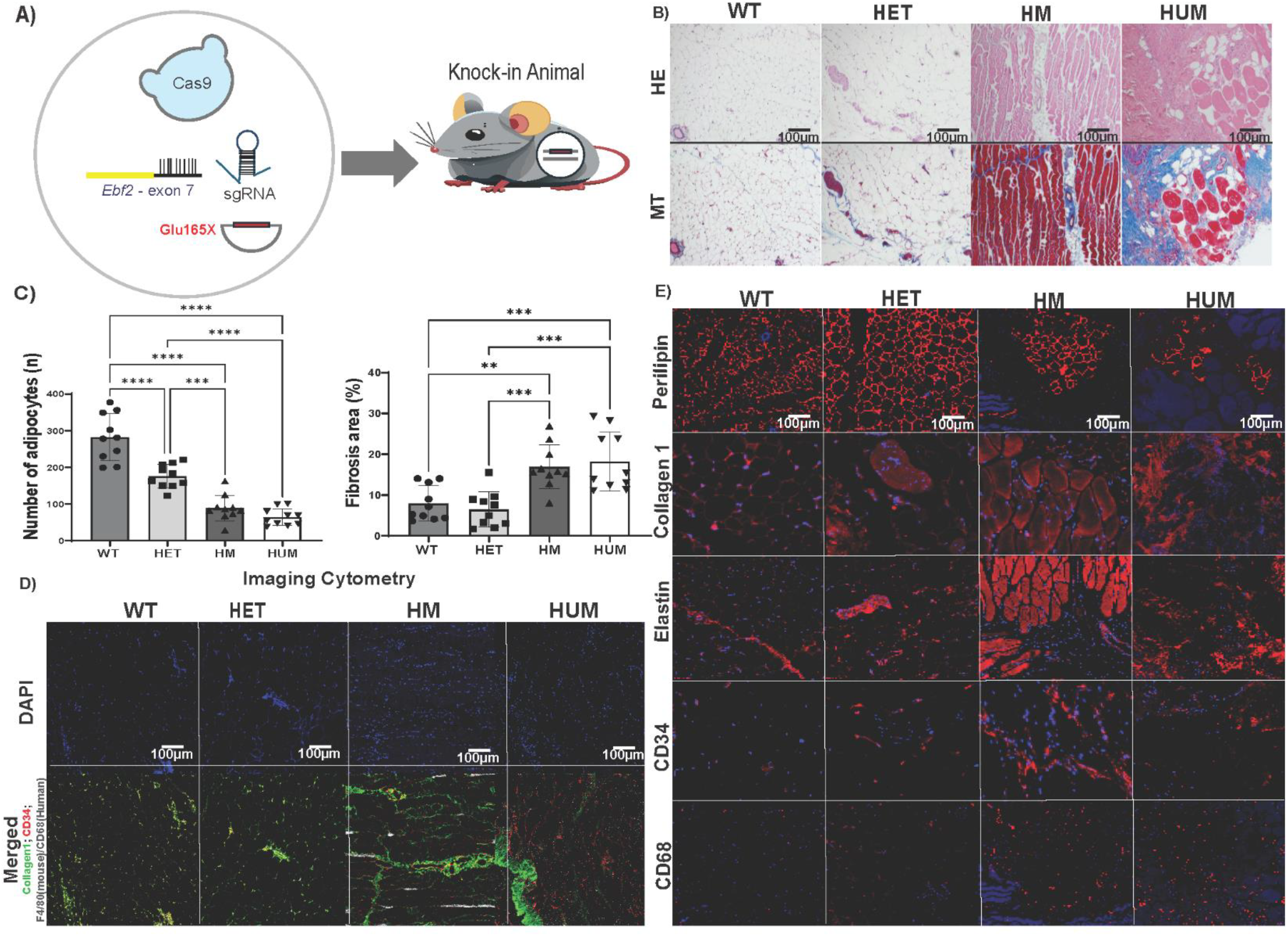
Inguinal WAT (IWAT) phenotype of *Ebf2mut* knock-in (KI) mice is similar to AT obtained from our patient. A) Knock-in of variant Glu165X in exon 7 with sgRNA and Cas9 ribonucleoprotein. B) Staining of white AT for Hematoxylin and eosin (HE), Masson Trichrome (MT) of IWAT of wild type (WT), heterozygous (HET), and homozygous (HM) E*bf2mut*-KI animals and back of the neck WAT depot of our patient (HUM, for human) (Scale=100µm). C) Number of adipocytes measured from the HE images (n=10 random images per group), and percentage of fibrosis measured from MT images (n=10 random images per group) (**p=0.01, ***p=0.001, ****p<0.001, data represents Mean±SEM, statistical test performed by One way ANOVA with Tukey’s multiple comparisons). D) Imaging Cytometry (CyTOF) obtained from IWAT samples from wild type (WT), heterozygous (HET), and homozygous (HM) E*bf2mut*-KI animals, and back of the neck depot of our patient (HUM, for human) samples (Scale=100µm), upper panels show DNA staining (Blue), lower panels show merged image of Collagen-1 (lime), CD34 (red) and F4-80 or mice samples or CD68 for human sample (white). Immunofluorescence staining for Perilipin, Collagen-1, Elastin, CD34 and CD68 in IWAT of wild type (WT), heterozygous (HET), and homozygous (HM) E*bf2mut*-KI animals, and human proband (HUM) samples (Scale=100µm).

## Discussion

In this report, we present a case of PLD caused by a nonsense variant of *EBF2* for the first time, adding a novel disease mechanism underlying a subtype of this condition. Although our case is a singleton, the *in vitro* and *in vivo* disease modeling provide evidence to establish the causality between the *EBF2* variant and the disease presentation. Our work also highlights the importance of deep tissue phenotyping in understanding the impact of observed variants in patients with PLD whose disease etiology remains elusive. In this case, the profound disruption in tissue architecture of the patient and striking resemblance of the knock-in model were crucial to drawing the causal link.

Our interest in this variant was spurred by the previous links of EBF2 to adipogenesis, as highlighted in the introduction. Moreover, EBF2 had emerged as a marker of a specific subset of visceral adipocytes in human AT single-cell studies^11^. The *in vivo* data was intriguing in demonstrating the profound impact of the variant in cultured cells, including human adipocytes, which supported the previous reports of knock-down experiments in 3T3-L1 cells^5^. However, our data also suggested a dominant negative effect of this variant inhibiting rescue conferred by full-length variant in cultured cells.

In addition to the phenotypic changes observed in cultured cells, the altered gene programs were in pathways that regulated ECM integrity and local inflammation. These findings supported the observations from the AT of our patient and the KI model in which we observed excess ECM accumulation and inflammatory myeloid cell infiltration in proximity to CD34^+^ cells. CD34^+^CD31^-^ mesenchymal stem cells differentiate into adipocytes during AT development and in obesity^15,16^. The increased number of CD34^+^ cells juxtaposed with excess ECM accumulation suggests the critical role played by EBF2 in regulating AT character and ECM homeostasis. EBF2 is a crucial transcription factor determining the fate of aging fibroblasts and osteoblasts ^6,17^. While we focused on adipogenesis *in vitro* and AT structure *in vivo*, the *EBF2* nonsense variant may exert tissue-specific fibrosis and inflammation in various tissues where *EBF2* is expressed. Indeed, previous studies of *Ebf1* and *Ebf2* knockout mice showed impaired development of kidneys and bones, respectively ^18-20^.

Our findings suggest that dysregulated ECM remodeling is coupled with aberrant cytokine expression, at least in the cultured cells, leading to activation of inflammation, which was observed in patient and KI model-derived tissues. We believe this progressive inflammation and dysregulated ECM remodeling may have played a major role in our patient’s metabolic deterioration. Indeed, the patient showed chronic insulin resistance and metabolic dysfunction-associated steatohepatitis (MASH) and later developed autoimmune diabetes and hepatitis^13,14^, suggesting the pathological coupling of lipodystrophy syndrome with immune dysregulation.

Although groundbreaking as a new cause of lipodystrophy, our study is limited by the observation of this rare variant in a single patient and our focus mainly on AT. However, EBF2 may have different important roles in other tissues, including bone formation and kidney podocyte differentiation^5,6,17^. EBF activity is also important for muscle development^21,22^. Our findings reveal more SMA+ cells and elastin in HM and HUM AT, indicating a need for further studies to clarify *EBF2*’s role in myofibroblast activity. Previously, disease links of EBF2 to Kallmann syndrome and ventral hernia have been reported^23,24^. Our patient had clinical findings of scoliosis, hand contractures, hypogonadotropic hypogonadism, disruption of the abdominal wall and renal mesangial expansion, and ECM accumulation, as described previously. The spectrum of clinical findings may all be consistent with EBF2 dysfunction, and future work is needed to define the precise pathophysiological abnormalities in other tissues.

In conclusion, our study identified a role for *EBF2* disruption in partial lipodystrophy for the first time.

We propose that this variant leads to aborted adipogenesis, accumulation of CD34^+^ cells, aberrant ECM deposition, and increased inflammatory myeloid cells in the AT. Though only speculative at this stage, the alteration in EBF2 may result in a novel pathophysiological mechanism that alters the integrity and function of other tissues.

## Data Availability

All data produced in the present study are available upon reasonable request to the authors

## Authors contributions

**MF-F** conducted experiments, analyzed data, collected patient data, organized figures, and drafted the manuscript. **DG** performed experiments. **AMR** contributed to experiments and participated in result discussions. **LP** analyzed genome sequencing data. **MO’L, HLR**, and **MSU** offered variant interpretation guidance and reviewed manuscript drafts. **AN** contributed to patient data collection. **PS** participated in result discussions and provided experimental guidance. **EAO** conceived the project, assembled the study team, formed all aspects of the collaborations, discussed and gathered data, designed, supervised and oversaw patient studies, provided clinical care to the patient, secured funding, critically reviewed data analyses, and contributed to manuscript writing. **THC** planned laboratory experiments, curated data, performed final analysis, and contributed to manuscript writing. All authors critically reviewed and approved the manuscript. EAO and THC jointly take responsibility for the integrity of the data.

## Acknowledgments

We are grateful to our patient and her mother for providing us with an important research question to pursue and for inspiring our translational pipeline. Clinical phenotyping was made possible by NIH grant 5R03DK074488 (Oral EA) and by Lipodystrophy Research Fund (Oral EA) established at the University of Michigan Medical School through philanthropic support (by Sopha Family, Baker family, Rosenblum family and White Point Foundation of Turkey). Infrastructure support has been enabled by NIH grants 2P30DK089503, 5P30DK020572, and UL1TR002240, as well as support from Caswell Diabetes Institute of Michigan Medicine. Sequencing and analysis were provided by the Broad Institute of MIT and Harvard Center for Mendelian Genomics (Broad CMG) and were funded by the NIH grant K23DK114551, National Human Genome Research Institute grants UM1HG008900 (with additional support from the National Eye Institute, and the National Heart, Lung and Blood Institute) and R01HG009141, and in part by Chan Zuckerberg Initiative grant DAF2019-199278 (https://doi.org/10.37921/236582yuakxy), an advised fund of Silicon Valley Community Foundation (funder DOI 10.13039/100014989). We thank The Vector Core of the University of Michigan (Thomas Lanigan and Roland Hilgart) for designing and preparing lentivirus constructs. We thank the Transgenic Animal Model Core of the University of Michigan (Thomas L. Saunders, Zachary T. Freeman, Elizabeth Hughes, Wanda Filipiak, Galina Gabrilina, and Honglai Zhang) for the design and production of the Ebf2 Glu165X transgenic mice.

## REFERENCES

1. Brown RJ, Araujo-Vilar D, Cheung PT, et al. The Diagnosis and Management of Lipodystrophy Syndromes: A Multi-Society Practice Guideline. J Clin Endocrinol Metab 2016;101(12):4500-4511. (In eng). DOI: 10.1210/jc.2016-2466.

2. Mosbah H, Akinci B, Araújo-Vilar D, et al. Proceedings of the annual meeting of the European Consortium of Lipodystrophies (ECLip) Cambridge, UK, 7-8 April 2022. Ann Endocrinol (Paris) 2022;83(6):461-468. (In eng). DOI: 10.1016/j.ando.2022.07.674.

3. Foss-Freitas MC, Akinci B, Luo Y, Stratton A, Oral EA. Diagnostic strategies and clinical management of lipodystrophy. Expert Rev Endocrinol Metab 2020;15(2):95-114. (In eng). DOI: 10.1080/17446651.2020.1735360.

4. Eldin AJ, Akinci B, da Rocha AM, et al. Cardiac phenotype in familial partial lipodystrophy. Clin Endocrinol (Oxf) 2021;94(6):1043-1053. (In eng). DOI: 10.1111/cen.14426.

5. Jimenez MA, Akerblad P, Sigvardsson M, Rosen ED. Critical role for Ebf1 and Ebf2 in the adipogenic transcriptional cascade. Mol Cell Biol 2007;27(2):743-57. (In eng). DOI: 10.1128/mcb.01557-06.

6. Kieslinger M, Folberth S, Dobreva G, et al. EBF2 regulates osteoblast-dependent differentiation of osteoclasts. Dev Cell 2005;9(6):757-67. (In eng). DOI: 10.1016/j.devcel.2005.10.009.

7. Rajakumari S, Wu J, Ishibashi J, et al. EBF2 determines and maintains brown adipocyte identity. Cell Metab 2013;17(4):562-74. (In eng). DOI: 10.1016/j.cmet.2013.01.015.

8. Seale P. Transcriptional Regulatory Circuits Controlling Brown Fat Development and Activation. Diabetes 2015;64(7):2369-75. (In eng). DOI: 10.2337/db15-0203.

9. Shao M, Zhang Q, Truong A, et al. ZFP423 controls EBF2 coactivator recruitment and PPARγ occupancy to determine the thermogenic plasticity of adipocytes. Genes Dev 2021;35(21-22):1461-1474. (In eng). DOI: 10.1101/gad.348780.121.

10. Shapira SN, Lim HW, Rajakumari S, et al. EBF2 transcriptionally regulates brown adipogenesis via the histone reader DPF3 and the BAF chromatin remodeling complex. Genes Dev 2017;31(7):660-673. (In eng). DOI: 10.1101/gad.294405.116.

11. Emont MP, Jacobs C, Essene AL, et al. A single-cell atlas of human and mouse white adipose tissue. Nature 2022;603(7903):926-933. (In eng). DOI: 10.1038/s41586-022-04518-2.

12. Agrawal S, Wang M, Klarqvist MDR, et al. Inherited basis of visceral, abdominal subcutaneous and gluteofemoral fat depots. Nature Communications 2022;13(1). DOI: 10.1038/s41467-022-30931-2.

13. Akinci B, Meral R, Rus D, et al. The complicated clinical course in a case of atypical lipodystrophy after development of neutralizing antibody to metreleptin: treatment with setmelanotide. Endocrinol Diabetes Metab Case Rep 2020;2020 (In eng). DOI: 10.1530/EDM-19-0139.

14. Altarejos JY, Pangilinan J, Podgrabinska S, et al. Preclinical, randomized phase 1, and compassionate use evaluation of REGN4461, a leptin receptor agonist antibody for leptin deficiency. Sci Transl Med 2023;15(723):eadd4897. (In eng). DOI: 10.1126/scitranslmed.add4897.

15. Lin CS, Xin ZC, Deng CH, Ning H, Lin G, Lue TF. Defining adipose tissue-derived stem cells in tissue and in culture. Histol Histopathol 2010;25(6):807-15. (In eng). DOI: 10.14670/HH-25.807.

16. Ravaud C, Esteve D, Villageois P, Bouloumie A, Dani C, Ladoux A. IER3 Promotes Expansion of Adipose Progenitor Cells in Response to Changes in Distinct Microenvironmental Effectors. Stem Cells 2015;33(8):2564-73. (In eng). DOI: 10.1002/stem.2016.

17. Nieminen-Pihala V, Rummukainen P, Wang F, Tarkkonen K, Ivaska KK, Kiviranta R. Age-Progressive and Gender-Dependent Bone Phenotype in Mice Lacking Both Ebf1 and Ebf2 in Prrx1-Expressing Mesenchymal Cells. Calcif Tissue Int 2022;110(6):746-758. (In eng). DOI: 10.1007/s00223-022-00951-7.

18. Fretz JA, Nelson T, Velazquez H, Xi Y, Moeckel GW, Horowitz MC. Early B-cell factor 1 is an essential transcription factor for postnatal glomerular maturation. Kidney Int 2014;85(5):1091-102. (In eng). DOI: 10.1038/ki.2013.433.

19. Nelson T, Velazquez H, Troiano N, Fretz JA. Early B Cell Factor 1 (EBF1) Regulates Glomerular Development by Controlling Mesangial Maturation and Consequently COX-2 Expression. J Am Soc Nephrol 2019;30(9):1559-1572. (In eng). DOI: 10.1681/ASN.2018070699.

20. Fretz JA, Nelson T, Xi Y, Adams DJ, Rosen CJ, Horowitz MC. Altered metabolism and lipodystrophy in the early B-cell factor 1-deficient mouse. Endocrinology 2010;151(4):1611-21. (In eng). DOI: 10.1210/en.2009-0987.

21. Green YS, Vetter ML. EBF proteins participate in transcriptional regulation of Xenopus muscle development. Dev Biol 2011;358(1):240-50. (In eng). DOI: 10.1016/j.ydbio.2011.07.034.

22. Jin S, Kim J, Willert T, et al. Ebf factors and MyoD cooperate to regulate muscle relaxation via Atp2a1. Nat Commun 2014;5:3793. (In eng). DOI: 10.1038/ncomms4793.

23. Jorgenson E, Makki N, Shen L, et al. A genome-wide association study identifies four novel susceptibility loci underlying inguinal hernia. Nat Commun 2015;6:10130. (In eng). DOI: 10.1038/ncomms10130.

24. Trarbach EB, Baptista MT, Garmes HM, Hackel C. Molecular analysis of KAL-1, GnRH-R, NELF and EBF2 genes in a series of Kallmann syndrome and normosmic hypogonadotropic hypogonadism patients. J Endocrinol 2005;187(3):361-8. (In eng). DOI: 10.1677/joe.1.06103.

