## Supplemental Appendix for "Early B-cell transcription factor-2 defect as a novel cause of lipodystrophy: disruption of the adipose tissue character and integrity"

### SUPPLEMENTARY METHODS

#### Patient Studies

**Ethics statement:**

Written consent was obtained from the patient, and all procedures involving the patient were approved by the Institutional Review Boards of Michigan Medicine and Massachusetts General Brigham and complied with the Declaration of Helsinki. All clinical characterization methods have also been published previously. [1]

**Genetic sequencing:**

PCR-free preparation of sample DNA (350 ng input at >2 ng/ul) is accomplished using Illumina HiSeq X Ten v2 chemistry. Libraries are sequenced to a mean target coverage of >30x.

Genome sequencing data was processed through a pipeline based on Picard, using base quality score recalibration and local realignment at known indels. The BWA aligner mapped reads to the human genome build 38. Single Nucleotide Variants (SNVs) and insertions/deletions (indels) are jointly called across all samples using the Genome Analysis Toolkit (GATK) HaplotypeCaller package version 4.0. Default filters were applied to SNV and indel calls using the GATK Variant Quality Score Recalibration (VQSR) approach. Annotation was performed using a Variant Effect Predictor (VEP). Lastly, the variant call set was uploaded to seqr for analysis.

1. **Cell culture**

3T3-L1 cells (ATCC, CL-173) and human subcutaneous preadipocytes (Cell Applications # 802s-05a) were cultured in Dulbecco's modified Eagle's medium (DMEM, Invitrogen) supplemented with 100 U/ml penicillin, 100 µg/ml streptomycin, 2 mM L-glutamine and 10% fetal bovine serum (Invitrogen, A3160402) at 37$^{o}$C and 5% $CO_2$.

Three days after reaching confluence, adipogenesis was induced by adding 0.25 µM dexamethasone, 1 µg/ml porcine insulin (all from Sigma Chemical Co., St. Louis, MO), and 0.5 mM IBMX for the 3T3-L1 cells and by adding or rosiglitazone (10 µM) for the human pre-adipocytes.

**Knockdown of murine *Ebf2* and lentiviral gene transfer of human *EBF2*, full-length and truncated variant.**

*siRNA*

Two different siRNA (Table 1) were purchased from ThermoFisher Scientific. 3T3-L1 cells were transfected with 5 nM siRNA targeting *EBF2* (MSS203753 and MSS203754) or control siRNA oligos using RNAi MAX (Invitrogen).

**Table 1:** Information on siRNA targeting *EBF2*

| | Cat No-Primer No | Primer name | Sequence |
|---|---|---|---|
| EBF2 #1 | 10620318-437913 A09 | Ebf2MSS203753 | (RNA)-CCA UAG CCU AUG AGG GAC AGA AUA A |
| | 10620319-437913 A06 | Ebf2MSS203753 | (RNA)-UUA UUC UGU CCC UCA UAG GCU AUG G |
| EBF2 #2 | 10620318-445249 B02 | Ebf2MSS203754 | (RNA)-CCC UCU ACG GCA CAC CAC ACA AUA A |
| | 10620319-445249 A10 | Ebf2MSS203754 | (RNA)-UUA UUG UGU GGU GUG CCG UAG AGG G |

*shRNA*

Five short hairpin RNAs (shRNAs) targeting distinct regions of the Ebf2 mRNA sequence were custom-designed by the Vector Core of the University of Michigan (Table 2). The pLKO.1 puro (Mission shRNA Sigma-Aldrich) lentiviral vector served as the delivery platform for these shRNAs.

**Table 2:** shRNA oligonucleotide sequences and targets for human and mouse *EBF2* gene

| Name | Alignment sequences | *EBF2* Target |
|---|---|---|
| shRNA Clone 1 | Hs. Not in construct as it targets the 3’ UTR.<br>However, it shares nuc 4-21 with mouse shRNA | GCAGCAATTCTTTGTTATGAA |
| shRNA Clone 2 | Hs.<br>GCATTAAATGAACCCACCATA<br>GCCTTAAATGAACCCACCATA | GCCTTAAATGAACCCACCATA |
| shRNA Clone 3 | HS.<br>TGGCTACAGCAATGTCCCCAT<br>CGGCTACAGCAATGTTCCTAT | CGGCTACAGCAATGTTCCTAT |
| shRNA Clone 4 | Hs.<br>GTTCATTTACACAGCATTAAA<br>GTTCATCTACACAGCCTTAAA | GTTCATCTACACAGCCTTAAA |
| shRNA Clone 5 | Hs.<br>GACAGAATAAGAATCCGGAAA<br>GACAGAATAAGAATCCCGAAA | GACAGAATAAGAATCCCGAAA |

**Lentivirus transduction**

Lentivirus containing five different clones of shRNA to knock down the murine Ebf2, along with either the full-length or truncated version of the human EBF2 sequence, were generated in the Vector Core at the University of Michigan. Transduction with lentivirus was conducted when cells reached 40-60% confluency, employing Polybrene at 4mcg/mL and 5% FBS overnight. Transduction efficiency was evaluated 48 hours post-incubation by observing the GFP signal. Cells containing shRNA were subsequently selected using Puromycin at 1mcg/mL.

**Assessment of EBF2 activity**

For analysis by luminescence measurement, HEK293 cells were transfected with pGL3-4XEBF2 enhancer SV40 Luc together with pRL-SV40 (Promega) per well using Lipofectamine 2000 (ThermoFisher Scientific). After 24 hours, cell lysate was collected in a Passive Lysis Buffer, followed by the detection of luciferase activity using the Dual Luciferase Assay System (Promega) and a GloMax microplate reader.

2. **Gene expression**

RNA extraction was performed using the RNeasy Mini Kit (Qiagen) and cDNA synthesis using IScript (Bio-Rad). Real-time quantitative PCR (qPCR) was carried out employing TaqMan probes for Pparg and Fabp4, recognized adipogenesis markers. Probe sequences are listed in Table 3. Quantitative PCR (qPCR) was conducted using the StepOnePlus Real-Time PCR system (Applied Biosystems, Foster City, CA).

**Table 3:** List of TaqMan probes used for qPCR.

| **Gene** | **Ref Seq Number** | **Probe Sequence** | **Primer Sequence** |
|---|---|---|---|
| *Pparg* | NM_011146 | 5`-/56-FAM/AGCTGACCC/ZEN/AATGGTTGCTGATTACA/3IABxFQ/-3’ | 5’CTGCTCCACACTATGAAGACAT-3’<br>5’-TGCAGGTTCTACTTTGATCGC-3’ |
| *Fabp4* | NM_024406 | 5`-/56- | 5’-AAATCACCGCAGACGACAG- |

| | | | |
|---|---|---|---|
| | | FAM/TGAAGAGCA/ZEN/TCATAACCCTAGATGGCG/3IABxFQ/-3’ | 3’ 5’-CCTTTCATAACACATTCCACCAC-3’ |
| *Ebf2* | NM_010095 | 5`-/56-FAM/ACATCCGCA/ZEN/ACACAAGCAGCATC/3IABxFQ/-3’ | 5’-GTCAGCATTTCAGAGTCCACA-3’ 5’-GGAGGTGCTGTAATTAGATTGCT-3’ |
| *Ebf1* | NM_007897 | 5`-/56-FAM/CTACACAGC/ZEN/ACTCAATGAACCCACCA/3IABxFQ/-3’ | 5’-TCGTACAAGTCCAAGCAGTTC-3’ 5’-GGAATGACCTTCTGTAACCTCTG-3’ |
| *Rplp0* | NM_007475 | 5`-/56-FAM/AGGCCCTGC/ZEN/ACTCTCGCTT/3IABxFQ/-3’ | 5’-TTATAACCCTGAAGTGCTCGAC-3’ 5’-CGCTTGTACCCATTGATGATG-3’ |
| *PPARg* | NM_005037 | 5'- /56-FAM/CCATGCTCT/ZEN/GGGTCAACAGGAGAAT/3IABkFQ/ -3' | 5'-TGTTATGGGTGAAACTCTGGG-3' 5'-TGTGAAGTGCTCATAGGCAG-3' |

| | | | |
|---|---|---|---|
| *FABP4* | NM_001442 | 5`-/56-FAM/CAGGAAAGT/ZEN/GGCTGGCATGGC/3IABxFQ/-3’ | 5’-ACTTGTCTCCAGTGAAAACTTTG-3’<br>5’-ATCACATCCCCATTCACACT-3’ |
| *EBF2* | NM_022659 | 5`-/56-FAM/TGCTTGTGT/ZEN/TGCGGATGTACCCTT/3IABxFQ/-3’ | 5’-GTCAGCATCTCAGAGTCAACA-3’<br>5’-CTGTAGCCATTCATACTGTTGC-3’ |
| *EBF1* | NM_024007 | 5`-/56-FAM/CCGCAGCCC/ZEN/TTTTGAGTATTACTTCCT/3IABxFQ/-3’ | 5’-CAGAGGTTACAGAAGGTCATTCC-3’<br>5’-CATCCCATACAGTGCTTCTACC-3’ |
| *RPLP0* | NM_001002 | 5`-/55UN/CCCTGTCTT/ZEN/CCCTGGGCATCAC/3IABxFQ/-3’ | 5’-TCGTCTTTAAACCCTGCGTG-3’<br>5’-TGTCTGCTCCCACAATGAAAC-3’ |

##### 3. Immunofluorescent staining

Cells were fixed with 4% paraformaldehyde, and inguinal fat pads were fixed in 1% paraformaldehyde at 4°C. Samples were then incubated in blocking solution (PBS with 5% BSA and 0.3% Triton X-100) and later with primary antibodies (diluted 1:100 in blocking solution) overnight at 4°C (Table 4). The samples were then incubated with Alexa Fluor–conjugated secondary antibodies (diluted 1:1000 in blocking solution) and DAPI (Invitrogen) diluted 1:10,000. Images were obtained using a fluorescent microscope (Olympus IX71, Olympus, Center Valley, PA) with a ×20 objective lens (NA 0.65) at 25°C.

Inguinal fat pads were fixed overnight in 1% paraformaldehyde at 4°C and processed in paraffin blocks. Unstained slides were incubated in a blocking solution (PBS with 5% BSA and 0.3% Triton X-100) for 1 hour at room temperature and later with primary antibodies (diluted 1:100 in blocking solution) overnight at 4°C (Table 4). After three washes with blocking solution, the tissues were incubated with Alexa Fluor–conjugated secondary antibodies (diluted 1:1000 in blocking solution) for 1 hour at room temperature, shielded from light.

Following three PBS washes, the tissues were stained with DAPI (Invitrogen) diluted 1:10,000 in water for 10 minutes, washed again three times with PBS, and imaged using a fluorescent microscope (Olympus IX71, Olympus, Center Valley, PA) equipped with a ×20 objective lens (NA 0.65) at 25°C.

**Table 4:** Immunoblot antibodies.

| Protein | Host | Dilution | Company | Catalog # |
|---|---|---|---|---|
| Perilipin | Rabbit | 1:100 | Cell signaling | 3470S |
| CD34 | Rabbit | 1:100 | Cell signaling | 26233S |
| Collagen 1 | Rabbit | 1:100 | Rockland | 600-401-103 |
| CD68 | Rat | 1:100 | BioRad | MCA1957 |
| Fibronectin | Rabbit | 1:100 | Sigma-Aldrich | F3648 |
| Elastin | Rabbit | 1:100 | Bioss | bs-1756R |
| SMA | Rabbit | 1:100 | Abcam | ab5694 |
| Bodipy | | 1:100 | Thermo Fisher | B3932 |

| | | | | |
|---|---|---|---|---|
| Donkey Anti-Rat IgG, Alexa flour 594 | Anti-Rat | 1:200 | Invitrogen | A-11007 |
| Goat anti-rabbit IgG, Alexa flour 488 | Anti-Rabbit | 1:200 | Invitrogen | A11070 |

**Image quantification for human and animal tissue:**

The intensity of perilipin, collagen 1, and CD34 was quantified with ImageJ version 1.6 after converting RGB TIFF to 8-bit binary images. Quantification of selected ROIs was performed using the "Analyze Particles" function in ImageJ, enabling measurement of parameters such as area and intensity. Using AdipoQ, an image J plugin, we quantified lipid droplet accumulation by determining the Bodipy positive area. We defined an adipogenic index by calculating the ratio of the Bodipy positive area and the DAPI positive area.

Digital images of the samples were acquired using a fluorescence microscope, maintaining consistent exposure settings across all images. Acquired images were imported into ImageJ software and adjusted for brightness and contrast to ensure optimal visualization of features while maintaining uniform adjustments across all images. ROIs corresponding to specific cell populations or tissue areas were outlined and selected using appropriate tools in ImageJ. Thresholding was applied to convert images into binary format, separating areas of interest from the background, with threshold levels adjusted to optimize separation while minimizing noise. Quantification of selected ROIs was performed using the "Analyze Particles" function in ImageJ, enabling measurement of parameters such as area and intensity. Settings such as minimum and maximum particle size were adjusted to exclude noise while capturing relevant features. Measurements, including mean fluorescence intensity and area covered, were recorded for each ROI. To quantify lipid droplets in cell culture, we used Bodipy. To visualize nuclei, we co-stained the cells with DAPI. We quantified lipid droplet accumulation using an Image J plugin by determining the Bodipy positive area. We defined an adipogenic index by calculating the ratio of the Bodipy positive area and the DAPI positive area.[2].

4. **Statistics**

The data are expressed as mean ± SEM unless stated otherwise and were subjected to analysis using the two-tailed Student t-test or ANOVA with Bonferroni's or Tukey's multiple comparison test. Statistical significance was determined at $P < 0.05$ or as specified in the figure legends. GraphPad Prism version 9.1.0 (221) was used for statistical analyses.

**SUPPLEMENTARY FIGURES LEGENDS:**

**Supplement Figure 1:** Presence of increased fibrotic tissue in the liver and presence of lamination, collagenous material accumulation, and fibrosis in the kidney tissue from our patient, which was performed for persistent massive proteinuria.

**Supplement Figure 2:** A) Two independent siRNA clones against *Ebf2* suppressed adipogenesis (siRNA#1 and siRNA#2). B) *Ebf2* and *Ebf1* expression in both siRNA-transfected cells (n=4, *p=0.02). C) *Pparg* and *Fabp4* expression assessed in adipocytes differentiated after si*Ebf2* #2 treatment. n=3. (*p=0.01, **p=0.001, ***p<0.001). D) *Ebf2* and *Ebf1* expression in 3T3-L1 cells transfected with five different lentiviral short hairpin RNA (shRNA) to knock down *Ebf2* showing a differential suppression against *Ebf2* relative to *Ebf1* (n=1). Data represents Mean±SEM. Statistical test performed by One way ANOVA with Tukey's multiple comparisons.

**Supplement Figure 3:** A) Mouse 3T3-L1 and Human SQ WAT-derived preadipocytes transduced with lentivirus control, EBF2-full, and EBF2-mut. Adipogenesis was induced, and lipid droplet accumulation was assessed (BODIPY= red, nucleus= blue). Representative data. Repeated more than four times. Adipocytes were quantified using ADIPOQ plug-in in Image J software, and cells transduced with lentivirus containing EBF2-mut show less BODIPY area (B) and adipogenic index (C) (n=3, **p=0.01, ***p<0.001). *FABP4* (D) and *PPARG* (E) expression in human preadipocytes treated with control, EBF2-full (WT) and EBF2-mut (Mut) and adipogenesis induced (Diff) (n=3, *p=0.04). Data represents Mean±SEM. Statistical tests were performed by One way ANOVA with Tukey's multiple comparisons.

**Supplement Figure 4:** A) Imaging Cytometry (CyTOF) obtained from IWAT samples from wild type (WT), heterozygous (HET), and homozygous (HM) E*bf2mut*-KI animals and from the back of the neck WAT depot of our patient (HUM, for human) samples showing DNA, Collagen-1, CD34, Ly-6G (mice samples) or CD15 (human sample), CD11b (mice samples) or CD11c (human sample), and F4/80 (mice samples) or CD14 (human sample) (Scale=100µm). B) Upper panels show immunofluorescence staining of adipose tissue for smooth muscle actin (SMA), and lower panels show Fibronectin in IWAT of wild type (WT), heterozygous (HET), and homozygous (HM) E*bf2mut*-KI animals, and the back of the neck WAT depot of our patient (HUM for human).

#### REFERENCE

1. Ajluni N, Meral R, Neidert AH, et al. Spectrum of disease associated with partial lipodystrophy: lessons from a trial cohort. Clin Endocrinol (Oxf) 2017;86(5):698-707. (In eng). DOI: 10.1111/cen.13311.
2. Sieckmann K, Winnerling N, Huebecker M, et al. AdipoQ-a simple, open-source software to quantify adipocyte morphology and function in tissues and in vitro. Mol Biol Cell 2022;33(12):br22. (In eng). DOI: 10.1091/mbc.E21-11-0592.

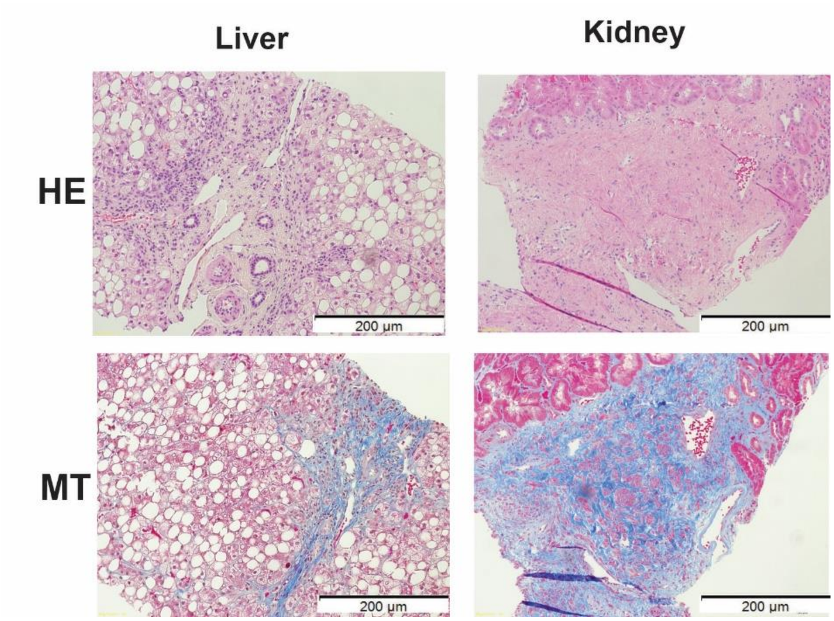

**Supplementary Figure 1**

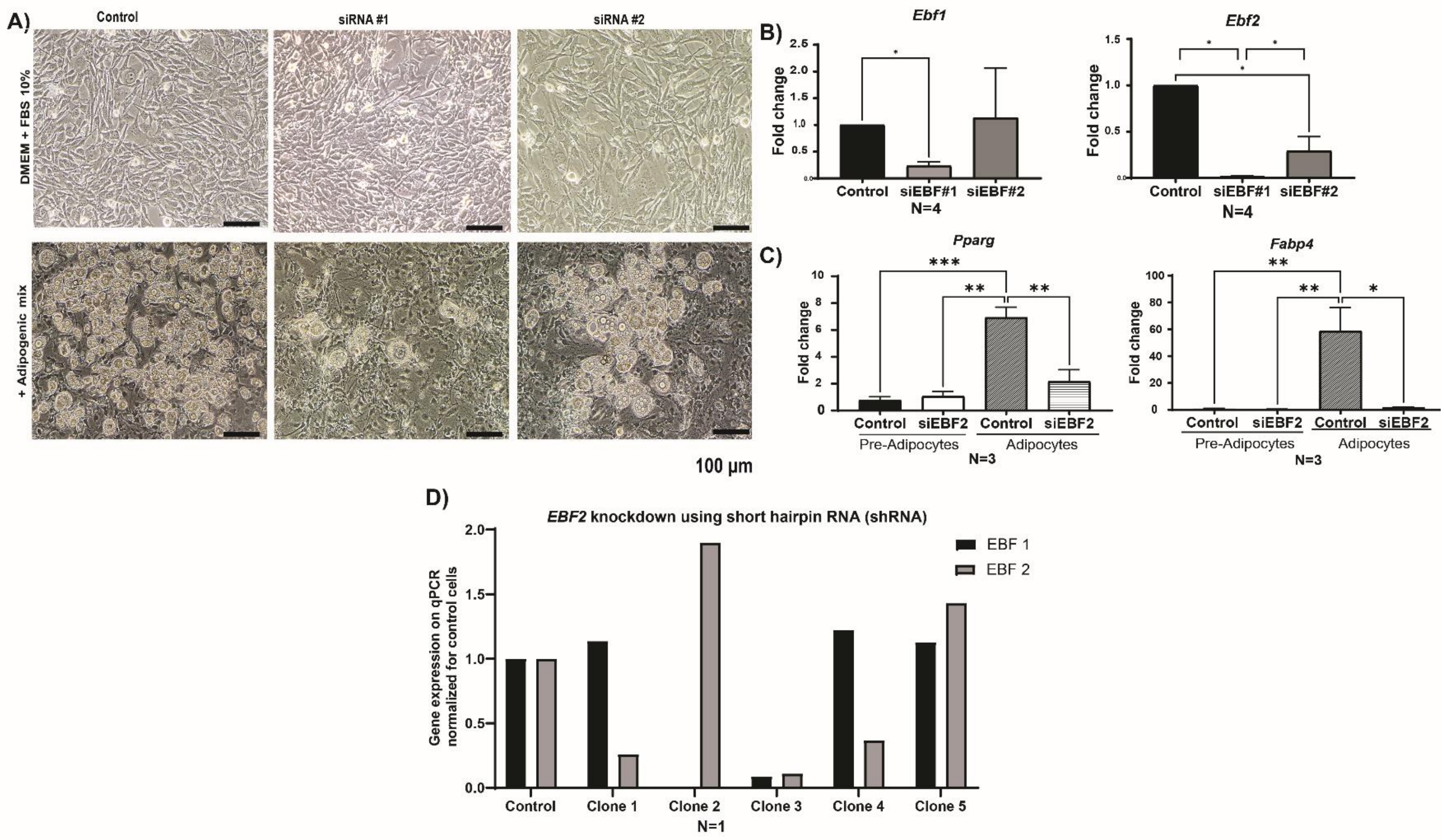

### Supplementary Figure 2

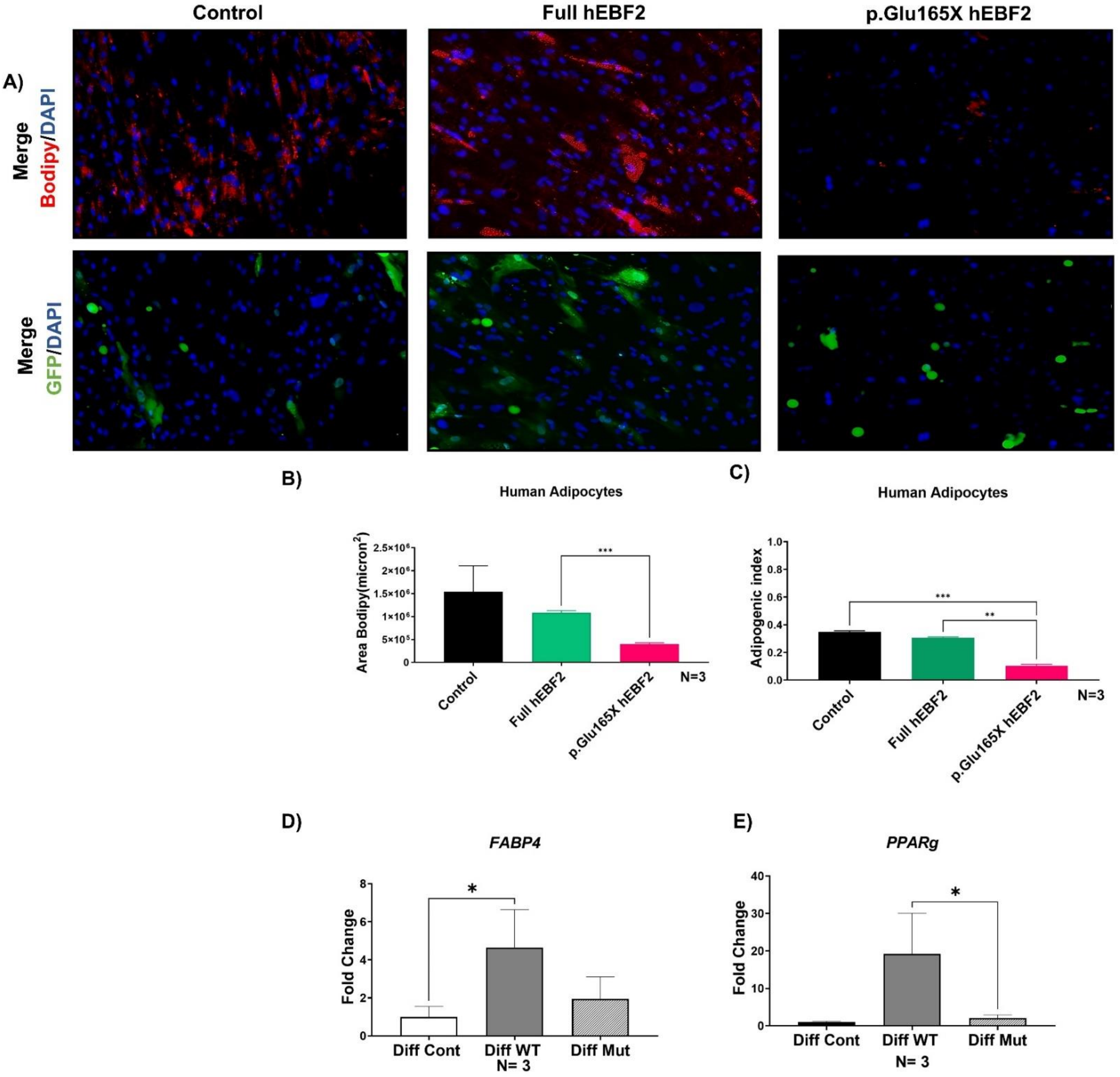

**Supplemental Figure 3**

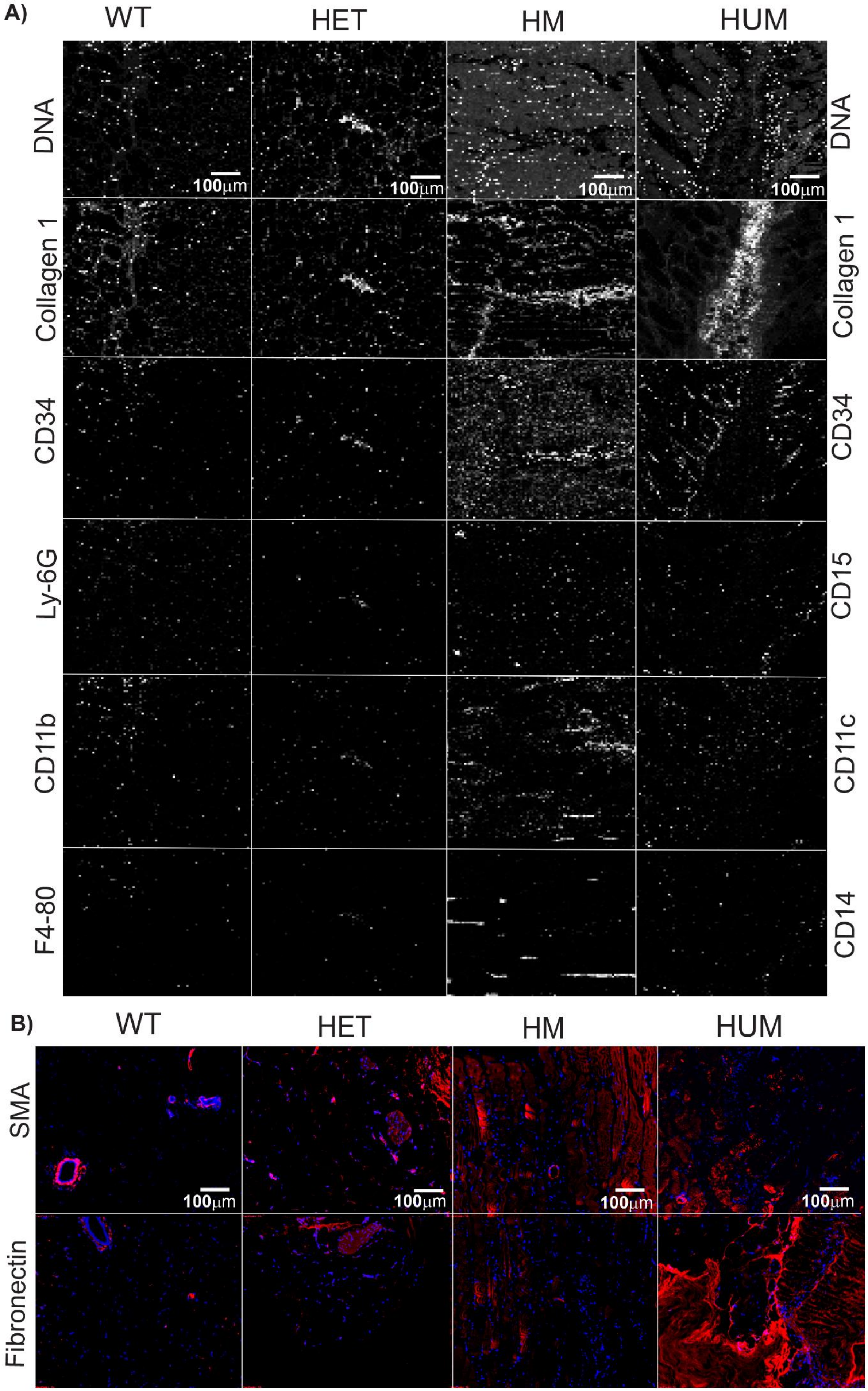

Supplemental Figure 4
